# School children’s oral health status, behaviours, and dental care utilization: the case of a northern Thailand elementary school

**DOI:** 10.1101/2024.02.25.24303350

**Authors:** Komkham Pattanaporn, Warathaya Kawong, Wassana Wongwanichwattana, Kulnan Chomphrom, Natchaphon Chakkathamanukul, Nalinporn Kamsiriphiman, Panithi Prasomphon, Pitchaya Saksurasap, Pansuk Nilniyom, Mario Brondani

## Abstract

**Background:** Although preventable, dental caries remain a serious global public health threat. There are a number of risk factors for the development of dental caries in the general population, and in elementary school children in Thailand in particular, especially if they are from suburban areas.

**Objective:** To identify the prevalence of dental caries in the form of decayed-missing-filled teeth for the permanent (DMFT) and primary (dmft) dentition, and their risk factors among children between 6 and 12 years of age attending a public school in Chiang Rai in northern Thailand.

**Methods:** A cross-sectional study was conducted among 6–12-year-old children attending public education at Thesaban 1 School, Chiang Saen District, Chiang Rai Province in Thailand. All school children from grades 1 to 6 were examined by trained dental students from Mae Fah Luang University’s School of Dentistry in December 2023. Children were clinically examined for DMFT/dmft while demographic information about the children was collected from school records. Children also responded to a standardized 17-item survey about their oral health care behaviours. Descriptive and inferential statistics were used. All tests were set at 95% CI and *p>*0.05.

**Results:** A total of 232 children (100% of eligible participants) were examined and 96 (40%) responded to the survey. Most children were Buddhists (n=186, 80.1%), and had at least one permanent tooth with sealant (n=198, 85.3%); 82.5% of the 232 children exhibited dental caries/extraction/fillings in primary teeth. The average dmft was 4.1 (1.8–8.8) and the average DMFT was 1.6 (0.1–3.3). Fifty children (21.5%) had their grandparents as their main caregiver. Children who brushed their teeth without fluoridated toothpaste were 1.99 times more likely to have at least 1 permanent tooth with dental decay than children who used fluoridated toothpaste (*p=*0.065).

**Conclusion:** In this study, a high percentage of dental caries was observed among children from suburban areas in the northernmost province of Thailand. Public health intervention and oral health promotion remain an immediate need among these children.

## INTRODUCTION

Chiang Rai is Thailand’s northernmost province, and part of the Golden Triangle region bordering Laos and Myanmar. Chiang Rai has the country’s largest number of remote hill tribes living in suburban and mountainous rural terrain who also experience social and health inequalities.^1^ In fact, almost 80% of the poorest individuals in Thailand reside in rural areas within agricultural households, while the distribution of poverty has been uneven in the South and Northeast—with Chiang Rai experiencing almost double the poverty rate at the national level.^2^ Nonetheless, an increasing number of children living in suburban areas attend primary schools supported by their own communities and the Thai government.^3^ Children attending school are generally the main focus of preventive and curative oral health services delivered by either dentists or dental nurses (a.k.a. dental therapists) in Thailand.^4^ Good oral health can be understood as the extent to which an individual of any age is able to adequately eat, relax, and socialize without active disease or discomfort.^5^ One of the main disruptors of good oral health is the presence of dental caries, particularly when accompanied by pain, discomfort while chewing, and social withdrawal. Although preventable, dental caries remain the most common disease worldwide and a serious public health threat, including in Thailand. Dental caries is a diet-mediated disease, but it also has a number of other risk factors for its development and progression in the general population, and in school children in particular, especially if they are from suburban and rural areas. Nationally and every 5 years, the Thailand Government implements its cross-sectional National Oral Health Survey (TNOHS). The TNOHS assesses the oral health of seven specific age groups: 3 years old, 5 years old, 12 years old, 15 years old, 35–44 years old, 60–74 years old, and 80–85 years old, and the impact of oral health related quality of life among 12-year-old children. Although the latest TNOHS (9^th^ cycle) included Chiang Rai province,^6^ it did not collect data from children between 6 and 11 years of age,^7^ which is an age group that can present with unique social and psychological factors associated with dental caries and dental care attendance.^8^ In addition, the TNOHS may not have equitably included suburban children from middle and low income neighbourhoods, further widening the lack of research on the oral health status and behaviours of Thailand’s middle to low income communities living in suburban areas.^1^ Therefore, this study aimed to identify the prevalence of dental caries in the form of diseased, missing, and filled teeth in primary (dmft) and permanent (DMFT) dentitions, and their risk factors among children between 6 and 12 years of age attending a public school in the Chiang Rai region. The study’s findings help to better understand the prevalence of dental decay and its related risk factors among suburban children in northern Thailand, with further implications for research related to health promotion and prevention, and overall quality of life within this demographic.

## METHODS

### Study procedures and sampling

The study received an ethical exemption from the Thesaban 1 School Ethics Board (#SC#2023-002). A cross-sectional study was conducted among 6–12-year-old children attending public education at Thesaban 1 School, Chiang Saen District, Chiang Rai Province in Thailand on December 24 and 25, 2023. All children from Grade 1 to 6 from that school were invited to participate upon verbal consent from their primary caregiver (parents, guardians). Although only a randomly selected sample of children participated in an interview survey, all children received intra-oral examination by trained senior dental students mentored by an experienced faculty member (KP) from Mae Fah Luang University’s School of Dentistry as part of a community dentistry course. A group of eight dental students (WK, WW, KC, NC, NK, PP, PS, PN) worked in pairs and collected all the DMFT/dmft data using an intra-oral plane mouth mirror, a World Health Organization periodontal probe, and a flash light source. The teeth were dried with gauze but not professionally cleaned before the examination. No X-rays were taken. The presence of sealant was recorded for each tooth.

The sample size for the intra-oral examinations was the total enrollment numbers of the school grades 1 to 6, while the sample size for the survey was determined considering a confidence level of 95% (α = 0.05), a relative degree of precision (d) of 10–15%, and a design effect (deff) of 2. Factoring in a dropout rate of 20%, the sample size for surveys was determined to be 96 children with an even distribution of males and females from each grade. This sample was drawn using the Simple Random Sampling method for the entire population. The results from these 96 interviews were extrapolated to the total sample given that these interviews were randomly selected and were representative of each grade by gender; we assumed the probability of the survey answers being consistent for the other students from their respective grade.

### Demographic information and surveys

Demographic information about the children’s religion (e.g., Buddhist, Christian, Muslim, and other), dental visits outside the school (e.g., yes/no), and primary caregiver (e.g., parent, grand parent, family member) and their employment status (e.g., farmer, government employee, general employee, other) were gathered from school records. Oral health care behaviours were gathered via a standardized 17-item survey applied to the child. The survey contained questions pertaining to the frequency of brushing (e.g., after meals/snacks, before bedtime, etc.), use of toothpaste (with or without fluoride), frequency of snacking (sugar-rich foods), and sugary beverage consumption (e.g., times per day and week), for example. The survey was administered by the senior dental students, who received answers from the children.

All data were anonymized and a unique code was attached to each child’s information for statistical purposes. Data analysis involved descriptive statistics, including mean and standard deviation, and inferential statistics using IBM SPSS Statistics for Windows (Version 19, IBM Corp., Armonk, N.Y., USA). The distribution of the dental caries index data (DMFT and dmft) was examined using average and range, while associations between variables were examined through the Chi-square test. All tests were set at 95% CI and *p*<0.05.

## RESULTS

The participating elementary school educates suburban neighbourhood children from middle to low income families and enrolls approximately 2% of hill tribe children from rural and underserved areas. From the 263 children attending the target school, 232 were between 6 and 12 years of age (Grades 1 to 6) and all were examined clinically, while 96 were randomly selected to participate in the survey (16 children per grade, 8 males and 8 females from each grade). Most children were Buddhists (n=186, 80.1%), had their parents as their main caregivers (n=161, 69.4%) and had at least one permanent tooth with sealant (n=198, 85.3%) (Table 1). Less than half of the children (46.5%, n=111) had dental caries in permanent teeth, while 75% (30 out of 40) of Grade 1 and 18.9% (7 out of 37) of Grade 6 children presented with no tooth decay on their permanent dentition (data not shown).

**Table 1.** Demographic characteristics, principal caregiver and their occupation, presence of sealants, consumption of snacks and sugary beverages between meals, brushing habits and visit to a dentist according to the grade level of 232 children examined at Thesaban 1 School in Chiang Rai, Thailand.

|  | **Grade 1**<br>**N=40(%)** | **Grade 2**<br>**N=48(%)** | **Grade 3**<br>**N=32(%)** | **Grade 4**<br>**N=41(%)** | **Grade 5**<br>**N=34(%)** | **Grade 6**<br>**N=37(%)** | **Total**<br>**N=232(%)** |
| --- | --- | --- | --- | --- | --- | --- | --- |
| **Gender** |  |  |  |  |  |  |  |
| Male | 21 (52.5) | 21 (43.7) | 12 (37.5) | 13 (31.7) | 11 (32.3) | 24 (64.8) | 102(43.9) |
| Female | 19 (47.5) | 27 (56.3) | 20 (62.5) | 28 (68.3) | 23 (67.7) | 13 (35.2) | 130(56.1) |
| **Religion** |  |  |  |  |  |  |  |
| Buddhist | 29 (72.5) | 31 (64.6) | 26 (81.2) | 36 (87.8) | 32 (94.1) | 32 (86.4) | 186(80.1) |
| Islam | 3 (07.5) | 3 (06.2) | 4 (12.5) | 5 (12.2) | 2 (05.9) | 5 (13.6) | 22(09.5) |
| Christian | 0 (00.0) | 3 (06.2) | 0 (00.0) | 0 (00.0) | 0 (00.0) | 0 (00.0) | 3(01.3) |
| Other<sup>Y</sup> | 8 (20.0) | 11 (23.0) | 2 (06.3) | 0 (00.0) | 0 (00.0) | 0 (00.0) | 21(09.1) |
| **Principal caregiver** |  |  |  |  |  |  |  |
| Parents | 34 (85.0) | 30 (62.5) | 30 (93.7) | 21 (51.2) | 23 (67.7) | 23 (62.1) | 161(69.4) |
| Grandparents | 6 (15.0) | 15 (31.3) | 0 (00.0) | 9 (21.9) | 6 (17.6) | 14 (37.9) | 50(21.5) |
| Family member | 0 (00.0) | 3 (06.2) | 0 (00.0) | 0 (00.0) | 3 (08.8) | 0 (00.0) | 6(02.6) |
| Other<sup>X</sup> | 0 (00.0) | 0 (00.0) | 2 (06.3) | 11 (26.9) | 2 (05.9) | 0 (00.0) | 15(06.5) |
| **Occupation of principal caregiver** |  |  |  |  |  |  |  |
| Farmer | 0 (00.0) | 0 (00.0) | 0 (00.0) | 5 (12.2) | 4 (11.8) | 2 (05.4) | 11(04.7) |
| Government | 3 (07.5) | 0 (00.0) | 4 (12.5) | 5 (12.2) | 4 (11.8) | 5 (13.5) | 21(09.1) |
| General employee | 16 (40.0) | 12 (25.0) | 8 (25.0) | 16 (39.0) | 5 (14.7) | 16 (43.3) | 73(31.4) |
| Other<sup>Z</sup> | 21 (52.5) | 36 (75.0) | 20 (62.5) | 15 (36.6) | 21 (61.7) | 14 (37.8) | 127(54.8) |
| **Sealant on permanent teeth\*** |  |  |  |  |  |  |  |
| Yes | 38 (95.0) | 44 (91.7) | 31 (96.9) | 37 (90.2) | 25 (73.6) | 23 (62.1) | 198(85.3) |
| No | 2 (05.0) | 4 (08.3) | 1 (03.1) | 4 (02.8) | 9 (26.4) | 14 (37.9) | 34(16.7) |
| **Children without dental decay on permanent teeth** | 32 (80.0) | 28 (58.3) | 14 (43.7) | 16 (39.0) | 11 (32.4) | 8 (21.6) | 108(46.5) |
| **Snacks between meals<sup>V</sup>** |  |  |  |  |  |  |  |
| 0 | 5 (12.5) | 0 (00.0) | 2 (06.3) | 0 (00.0) | 2 (05.9) | 0 (00.0) | 9(03.9) |
| 1-3 days | 21 (52.5) | 27 (56.3) | 24 (75.0) | 28 (68.3) | 21 (61.7) | 21 (56.7) | 143(61.6) |
| 4-6 days | 10 (25.0) | 18 (37.5) | 2 (06.3) | 10 (24.4) | 6 (17.7) | 12 (32.5) | 58(25.0) |
| Everyday | 2 (05.0) | 4 (08.2) | 4 (12.4) | 3 (07.3) | 5 (14.7) | 4 (10.8) | 22(09.5) |
| **Sugary beverages between meals<sup>V</sup>** |  |  |  |  |  |  |  |
| **None** | 11 (27.5) | 12 (25.0) | 2 (06.2) | 3 (07.3) | 0 (00.0) | 7 (18.9) | **35(15.1)** |
| **1-2 daily** | 18 (45.0) | 30 (62.6) | 24 (75.0) | 30 (73.1) | 19 (55.9) | 19 (51.2) | **140(60.3)** |
| **2-3 daily** | 8 (20.0) | 3 (06.2) | 4 (12.6) | 3 (07.3) | 12 (35.3) | 7 (18.9) | **37(15.9)** |
| **>3 daily** | 3 (07.5) | 3 (06.2) | 2 (06.2) | 5 (12.3) | 3 (08.8) | 4 (11.0) | **20(08.6)** |
| **Brush with**<br>**fluoridated**<br>**toothpaste<sup>T</sup>** | 26 (65.0) | 33 (68.7) | 32 (100) | 38 (92.7) | 32 (94.1) | 32 (86.5) | **193(83.2)** |
| **Daily brushing**<br>**habits<sup>~</sup>** |  |  |  |  |  |  |  |
| **After waking up** | 34 (85.0) | 42 (87.5) | 25 (78.2) | 33 (80.5) | 32 (94.1) | 23 (62.2) | **189(81.5)** |
| **After breakfast** | 8 (20.0) | 3 (06.2) | 4 (12.5) | 3 (07.3) | 0 (00.0) | 0 (00.0) | **18(07.8)** |
| **After lunch** | 5 (12.5) | 0 (00.0) | 10 (31.3) | 0 (00.0) | 0 (00.0) | 0 (00.0) | **15(06.5)** |
| **Brush before going**<br>**to bed (no food**<br>**after)** |  |  |  |  |  |  |  |
| **Daily** | 24 (60.0) | 23 (47.9) | 24 (75.0) | 18 (43.9) | 23 (67.6) | 23 (62.2) | **135(58.2)** |
| **Occasionally** | 8 (20.0) | 16 (33.3) | 6 (18.7) | 13 (31.7) | 11 (32.4) | 14 (13.8) | **68(29.3)** |
| **Never** | 8 (20.0) | 9 (18.8) | 2 (06.3) | 10 (24.4) | 0 (00.0) | 0 (00.0) | **29(12.5)** |
| **Seen a dentist**<br>**outside school<sup>H</sup>** |  |  |  |  |  |  |  |
| **Yes** | 35 (87.5) | 27 (56.3) | 22 (68.8) | 23 (56.1) | 28 (82.3) | 26 (70.2) | **161(69.4)** |
| **No** | 5 (12.5) | 21 (43.7) | 10 (31.2) | 18 (43.9) | 6 (17.7) | 11 (29.8) | **71(30.6)** |

<sup>Y</sup> None, or not informed, or other.
<sup>X</sup> Non-family related, or a community member.
<sup>/</sup> Including not informed, part-time work, or self-employed.
<sup>\*</sup> At least one permanent teeth with sealant.
<sup>v</sup> After any meals, per day.
<sup>T</sup> Brushing alone, under supervision or brushed by somebody else.
<sup>~</sup> Numbers and percentages overlap as the children might have different daily brushing habits, e.g., some might brush only after waking up, or after waking up and after lunch, or after every meal, and so on.
<sup>H</sup> For any reason.

The average decay-missing-filed primary teeth (dmft) was 4.1 (1.0–8.8) and the average for permanent teeth (DMFT) was 1.6 (0.1–3.3) (Table 2). A total of 80 children (34.55%) snacked 4 or more times a day and 39 children (16.8%) did not use fluoridated toothpaste (Table 1). Children who had their grandparents as principal caregivers were 2.6 times (*p=*0.093) and 2.37 times (*p=*0.133) more likely to snack in between meals more than 4 days a week and to consume sugary beverages more than 3 times a day, respectively, than children with parents or other family members as their principal caregivers; only the first odds ratio was statistically significant (Table 3).

**Table 2.** Average DMFT/dmft (and range) according to the grade level of 232 children examined at Thesaban 1 School in Chiang Rai, Thailand.

|  | Grade 1 | Grade 2 | Grade 3 | Grade 4 | Grade 5 | Grade 6 | Total |
| --- | --- | --- | --- | --- | --- | --- | --- |
| **DMFT** | 0.2 (0.1-0.7) | 1.2 (0.9-1.5) | 1.5 (1.1-1.9) | 1.5 (1.0-1.9) | 2.3 (1.9-2.6) | 3.0 (2.6-3.3) | **1.6 (0.1-3.3)** |
| **dmft** | 5.7 (4.1-8.8) | 5.4 (3.9-7.8) | 3.4 (1.9-4.2) | 2.6 (2.2-2.8) | 1.3 (1.0-1.5) | 1.5 (1.1-1.7) | **4.1 (1.0-8.8)** |

**Table 3.** Odds Ratio (OR), Confidence Interval (Cl) and p values (P) of various associations.

|  | Principal caregiver:<br>Grandparents | Snacking ><br>4 days per week | Sugary beverages ><br>3 times/day | Brushing without<br>fluoridated toothpaste | Brushing daily before bed | Decay in at least 1 permanent tooth |
| --- | --- | --- | --- | --- | --- | --- |
|  | OR CI P | OR CI P | OR CI P | OR CI P | OR CI P | OR CI P |
| Snacking > 4 days per week | 2.60<br>0.850-7.978<br>0.069 | N/A | N/A | N/A | N/A | 1.11<br>0.653-1.889<br>0.698 |
| Sugary beverages > 3 times/day | 2.37<br>0.767-7.358<br>0.133 | N/A | N/A | N/A | N/A | N/A |
| Brushing with fluoridated toothpaste | N/A | N/A | N/A | N/A | 12.63<br>2.976-53.668<br><0.001 | N/A |
| Decay in at least 1 permanent tooth | N/A | 1.11<br>0.653-1.889<br>0.698 | 0.12<br>0.065-0.242<br><0.001 | 1.99<br>0.956-4.145<br>0.065 | N/A | N/A |

Children who brushed their teeth (or had their teeth brushed for them) without fluoridated toothpaste were 1.99 times more likely to have at least 1 permanent tooth with dental decay than children who used fluoridated toothpaste (*p*=0.065). Children who brushed their teeth (or had their teeth brushed for them) every night before going to bed and did not consume any foods afterwards were 12.63 times more likely to have used fluoridated toothpaste than children who did not brush (or have their teeth brushed for them) every night before going to bed (*p*<0.01). Lastly, children with decay in at least one permanent tooth were 1.11 times more likely to consume snacks in between meals more than 4 days a week than children without decay on permanent teeth (*p*=0.698).

## DISCUSSION

The noted prevalence of dental caries among the children studied herein corroborates other studies from Thailand^9^ and internationally;^10,11^ although, its cross-sectional design does not allow us to make any inferences about whether the prevalence is decreasing locally, as seen globally.^12,13^ However, the findings of our study seem to be a bit higher for DMFT and a bit lower for dmft in comparison to other studies. For example, a previous inquiry conducted in Southern Thailand^14^ found that at age 6 (Grade 1), children had a mean dmft of 8.1, while ours have a mean dmft of 5.7. Likewise, that study showed an average DMFT at age 12 (Grade 6) of 2.4, while ours was 3.5. Our DMFT results at age 12 seem to also be higher than the national mean of 1.4 for Thai children.^15^ In fact, since that study in Southern Thailand, published in 2001, the oral health status of Thai school-age children has improved,^13^ although we could not corroborate that statement.

On the other hand, the fact that no more than 34% of the children (n=80) snacked in between meals more than 4 days in a week, and no more than 25% (n=57) consumed sugary beverages at least twice a day, might help to explain the finding that almost half of the children (46%, n=108) did not have any decay on permanent teeth. That is, perhaps our cohort’s snacking and sugary drink consumption was different than that found in other studies,^16,17^ although the odds ratio of 0.12 (*p*<0.001) of children consuming sugary beverages more than three times a day and having at least 1 permanent tooth with decay may indicate a decreased occurrence of decay, which is counterintuitive. However, the high consumption of sugar-rich beverages and snacks by some of the other children is still worrisome given the association with a high prevalence of dental decay in their teeth, as observed in this study and discussed by Skafida and Chambers.^18^ Moreover, the potential implication of sugar consumption on the development of inflammatory conditions such as periodontal disease^19^ (not measured in this study) calls for further reduction of daily sugar consumption overall.^20^

Despite the fact that only 16.8% of the children (n=39) did not use fluoridated toothpaste to brush their teeth/have them brushed, we strongly recommend the use of fluoridated toothpaste given its benefits in preventing caries.^21^ Further studies should explore whether or not the high consumption of snacks and sugary drinks in between meals would negate the benefits of fluoride, as suggested by Gage and colleagues^16^ and Sheiham and James.^20^ We were not able to collect any information about the consumption of fluoridated water by the participating children.

The presence of sealants in virtually all children (98% of them, with at least one permanent tooth with pit and fissure sealant) attests to the availability of dental care and the focus on prevention by the Thai government.^4^ In fact, the pit and fissure sealants could have been administered at school or in a dental office given the relatively high number of children visiting dentists outside of school. As these sealants can effectively prevent dental caries by forming a protective barrier that reduces bacterial growth and trapped food,^22^ we urge the widespread use of this preventive measure in community settings.

There is some evidence that grandparents tend to offer more snacks and sugary beverages to children compared to their parents,^23,24^ as we found in our study. Given the association between sugar consumption and caries development, it was not surprising to find that children cared for mostly by their grandparents had the highest consumption of such food items and the highest diseased-missing-filled teeth compared to the children cared for mostly by their parents (data not shown), supporting the findings from Morita and colleagues.^25^ However, the extent to which dental decay impacts the oral health-related quality of life of the children we examined remains unknown.

Although the TNOHS has a large sample size, it only includes 3-, 5-, and 12-year-old children,^6,7^ while many other Thai studies have focused on dental caries among preschool children younger than 6 years of age.^26,27^ Our study adds to the body of literature by including children between the ages of 6 and 12 with oral hygiene and eating habits associated with dental caries,^8^ and who are from suburban and middle to low income neighbourhoods.^1^ Lastly, the engagement of undergraduate dental students in studies like this, as part of their training, attests to the importance of community-driven dental education that aims at graduating socially responsible^28^ and reflective oral health care providers.^29^

Despite the participating school’s children receiving routine free-of-charge examinations, prevention, and treatment by oral health care providers under Universal Health Coverage,^30^ the somewhat high mean of dental caries, fillings, and extractions is still a concern, as we found in this study. Our findings may suggest that inadequate oral health care practices may be contributing, at least in part, to the prevalence of dental caries, fillings, and extractions among the target primary school students. Initiatives should prioritize interventions aimed at improving oral health care behaviours rather than merely increasing access to dental services.

Despite its findings, this study has limitations. The cross-sectional design prevents the establishment of causation for the associations reported herein. The focus on only one school prevents generalization despite the sample size being the total number of Grade 1–6 children in that participating school. Although the intra-oral examinations were conducted by trained senior dental students in their last year of education, the lack of intra- and inter-rater calibration might have jeopardized the accuracy of the results.

The survey used standard and easy to understand questions, but it was not validated or piloted prior to the study. Moreover, as the children themselves responded to the survey their answers might not have been accurate, even though research has shown adequate understanding and validity of reports provided by a child about their oral health at age 6 and older.^31,32^ In addition, there might have been many confounding factors influencing the associations we found herein that were not considered, including principal caregivers educational levels, family income, access to fresh fruits and vegetables, housing conditions, among others.

## CONCLUSIONS

In our study, a high prevalence of dental caries was observed among children in suburban areas in the northernmost province of Thailand, some of whom were likely facing many inequities. The number of children with dental caries, fillings, and extractions in the permanent dentition is still a concern. Further studies are needed to investigate the influence of confounding factors such as access to fresh fruits and vegetables on the associations we found. Public health intervention and oral health promotion remain an immediate necessity among these children.

## Data Availability

The data underlying the results presented in the study are partially available from the first author, Komkham Pattanaporn at

## ABBREVIATIONS

DMFT/dmft: Diseased, Missing, Filled Teeth
TNOHS: Thailand National Oral Health Survey

## DECLARATIONS

### Ethical approval and consent to participate

The study received an ethical exemption from the Thesaban 1 School Ethics Board (#SC#2023-002).

### Consent for publication

Not applicable

### Availability of data and materials

Full data cannot be shared publicly in its original Thai language and belongs to the creators. Partial data are available from the first author (KP).

### Authors’ contributions

KP contributed to the study conception, design, and data acquisition, and translated the data from Thai to English. WK, WW, KC, NC, NK, PP, PS, and PN collected the data and revised the manuscript. MB drafted the manuscript, developed the tables, interpreted the findings, and organized the references.

## ACKNOWLEDGMENTS

The authors would like to acknowledge all the school children and their parents and caregivers who made this study possible. A special thanks goes to the school staff who welcomed us and provided the needed support on site. This study received funds from the School of Dentistry, Mae Fah Luang University.

